# Genetic Variants that Modulate Alzheimer’s Disease Risk Deregulate Protein-Protein Correlations in the Gyrus Temporalis Medius

**DOI:** 10.1101/2025.02.11.25322052

**Authors:** Gerard A. Bouland, Niccolò Tesi, Meng Zhang, Andrea B. Ganz, Marc Hulsman, Sven van der Lee, Marieke Graat, Annemieke Rozemuller, Martijn Huisman, Natasja M. van Schoor, Wiesje M. van der Flier, Jeroen Hoozemans, August B Smit, Marcel J.T. Reinders, Henne Holstege

## Abstract

We conducted a comprehensive protein quantitative trait loci (pQTL) analysis on a unique set of Gyrus Temporalis Medius (GTM) samples obtained from 88 Alzheimer’s Disease (AD) patients, 53 non-demented individuals, and 49 cognitively healthy centenarians. This investigation revealed 8,081 genetic variants associated with the abundance of 227 proteins, including several novel variant-protein links not identified in a prior pQTL study of the prefrontal cortex or expression QTL (eQTL) analysis across 12 brain regions (GTEx). Among all the AD risk variants tested for possible pQTL effects, only rs429358-T (which encodes the APOE4 allele) was significantly linked to higher APOE levels in the GTM, potentially explaining why this region is particularly prone to AD pathology. Further, through differential correlation analysis we identified AD risk variants linked to altered protein-protein correlations, specifically rs9381040 in TREML2, rs34173062 in SHARPIN, and rs11218343 near SORL1. Notably, DDX17 appears to play a protective role in individuals with the rs9381040-T/T genotype by tightly regulating synuclein levels. Collectively, these findings demonstrate that AD risk variants disrupt protein–protein interactions, highlighting a genetic basis for the coordinated modulation of protein networks in AD.

## INTRODUCTION

Alzheimer’s Disease (AD) is a progressive disease marked by the loss of cognitive functions and autonomy, eventually leading to death^1^. Numerous genome wide association studies (GWASs) have been conducted to identify genetic modifiers of AD risk, including attempts to understand their role in AD etiology^2–4^. However, elucidating the mechanistic pathways through which these genetic loci influence AD-related processes is difficult. Many risk variants are common, have a small effect size on AD, and are located in non-coding and intergenic regions. These variants merely act as ‘markers’ of haplotypes, genetic regions averaging 300 kb that are inherited across generations^5,6^. These haplotypes typically include at least one genetic factor that increases disease risk, possibly by affecting the expression of one or more genes in the risk locus. Therefore, while GWAS ‘risk’ variants are unlikely to be directly causative, they are likely in (partial) linkage with the causal variant(s).

It is thus reasonable to investigate whether a risk locus is an expression quantitative trait locus (eQTL), meaning it is associated with changes in the expression of messenger RNAs (mRNAs)^7^. However, while mRNAs encode proteins, they often exhibit abundance profiles distinct from mRNAs. Consequently, many eQTLs show limited correspondence at the protein level and do not correspond to protein-QTLs (pQTLs)^8^. Since proteins are prominent determinants of the cell’s biology, understanding how AD risk alleles associate with protein expression could therefore help to reveal the genetic (de)regulation associated with these. Additionally, causative variants might have downstream effects that are missed when only investigating eQTLs/pQTLs. For instance, while a causative missense variant might not alter mRNA levels, the change in the amino acid sequence of a protein can lead to a significant change in protein function and interactions with other proteins, potentially altering various biological pathways. Assuming that co-expressed proteins are functionally related, allele-specific correlation patterns of protein abundance might indicate a unique regulatory state of biological pathways in response to an allele.

Based on this, we hypothesized that AD-associated alleles may have downstream functional consequences detectable through comparisons of protein co-expression between carriers and non-carriers of risk alleles. To explore this, we used a unique collection of brains from 88 AD patients, 53 non-demented individuals, and 49 cognitively healthy centenarians. We focused on the Gyrus Temporalis Medius (GTM), an area known for its vulnerability to AD-related neuropathological changes^9,10^. First we performed an extensive pQTL analysis to determine whether the abundance of measured proteins correlated with the presence of specific genetic variants (Fig. 1a). We then compared our findings with previously identified brain pQTLs^8^ and eQTLs in twelve brain regions from the GTEx database^7^, and evaluated each pQTL variant for its association with AD risk. Finally, to assess the impact of AD risk variants identified in a previous GWAS^2^ on protein correlation-structures, we performed a differential correlation QTL (dcQTL) analysis (Fig. 1b), examining changes in protein co-expression structures among carriers versus non-carriers of these alleles.

**Figure 1.**
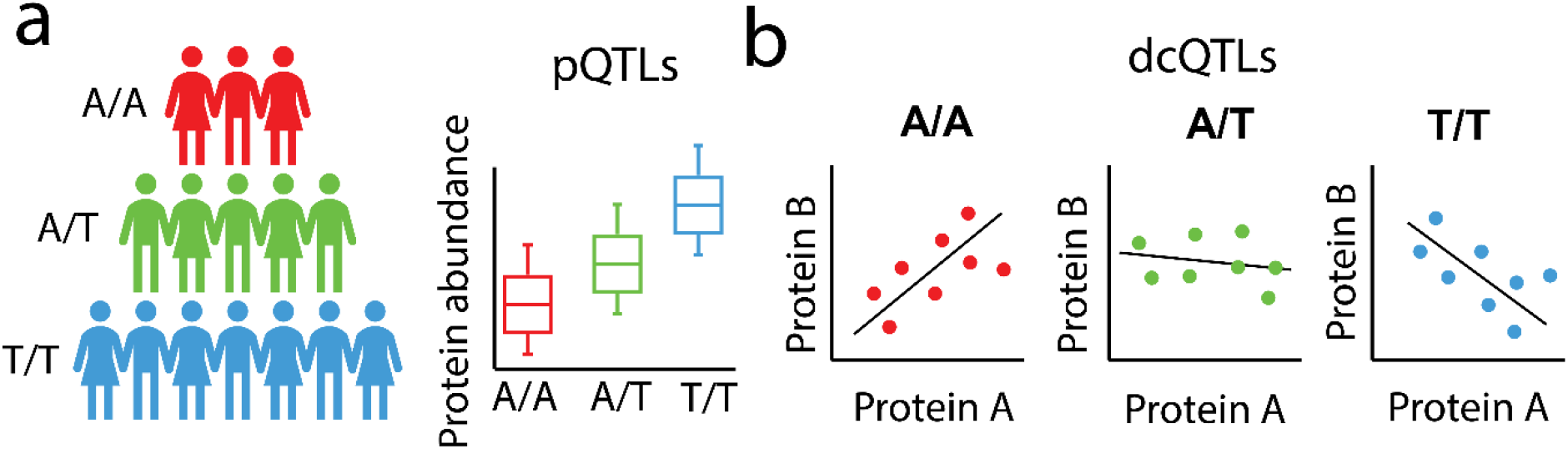
Overview of the pQTL and dcQTL analyses within this study. a) Schematic representation of a pQTL analysis. Individuals are grouped according to the genotypes of a genetic variant. A pQTL is identified when the expression of a protein is significantly (linearly) associated with the genotype. b) Schematic overview of the dcQTL analysis, identifying changes in co-expression relative to genotype. Individuals are grouped based on the genotypes of a genetic variant and pairs of proteins are identified whose correlation within each genotype differs between genotypes (P_FDR_ ≤ 0.05).

### Demographics

Protein abudance data from the Gyrus Temporalis Medius (GTM) were obtained for 190 individuals (mean age 86.8 ± 13.8, 73.7% female). Following quality control on genetic data, 6,607 individuals (mean age 68.4 ± 15.8, 53.7% female) remained in the study. Of these, 140 participants (mean age 91.0 ± 14.2, 74.3% female) had both proteomics and genetic data available (Table 1).

**Table 1:** Population characteristics of individuals for whom genotyping and protein data were available, including the intersection. AD = Alzheimer’s Disease cases, ND = Non-demented controls, CEN = centenarians.

|  | genotyping data |  |  | Protein data |  |  | intersection |  |  |
| --- | --- | --- | --- | --- | --- | --- | --- | --- | --- |
| Number of individuals | 6,607 |  |  | 190 |  |  | 140 |  |  |
| Females (%) | 3549 (53.7) |  |  | 140 (73.7) |  |  | 104 (74.3) |  |  |
| Diagnosis | AD | ND | CEN | AD | ND | CEN | AD | ND | CEN |
| N | 2,416 | 3,848 | 343 | 88 | 53 | 49 | 67 | 27 | 46 |
| Age ($\sigma$) | 70.2<br>(10.5) | 61.1<br>(14.8) | 101.0<br>(2.5) | 81.2<br>(11.2) | 81.1<br>(12.0) | 103.0<br>(2.3) | 79.6<br>(12.2) | 83.4<br>(8.7) | 103.1<br>(2.2) |

### Genetic modulation of protein abundance is largely independent from modulation of its corresponding mRNA in the Gyrus Temporalis Medialis

A pQTL analysis (Fig. 1a, Supplementary **Table 1**) including 140 individuals for which both genetic and proteomics data was available, was conducted (Table 1). pQTLs were identified using linear regression models, adjusting for estimated cell type composition (neurons, microglia/macrophages, and oligodendrocytes) and population substructure using the first five principal components (Supplementary file: ‘*pQTL linear regression model’*). A total of 3,427 proteins were tested for association with genetic cis-variants lying within 250 Kbp of the transcription start site (TSS). We identified 8,081 variants significantly associated with the abundance of 227 proteins in the GTM (P_FDR_ ≤ 0.05, Fig. 2a). Of these, 5,331 (∼66%) are new associations involving 150 proteins, while 2,750 variants (34%), were associated with 77 proteins, overlapping with pQTLs previously identified in a QTL study of the dorsolateral prefrontal cortex^8^. While some overlap in genetic associations across brain regions is anticipated, distinct protein abundance profiles in each region^11^ also lead to unique findings, which is consistent with the new and overlapping pQTL signals observed in the GTM.

**Figure 2.**
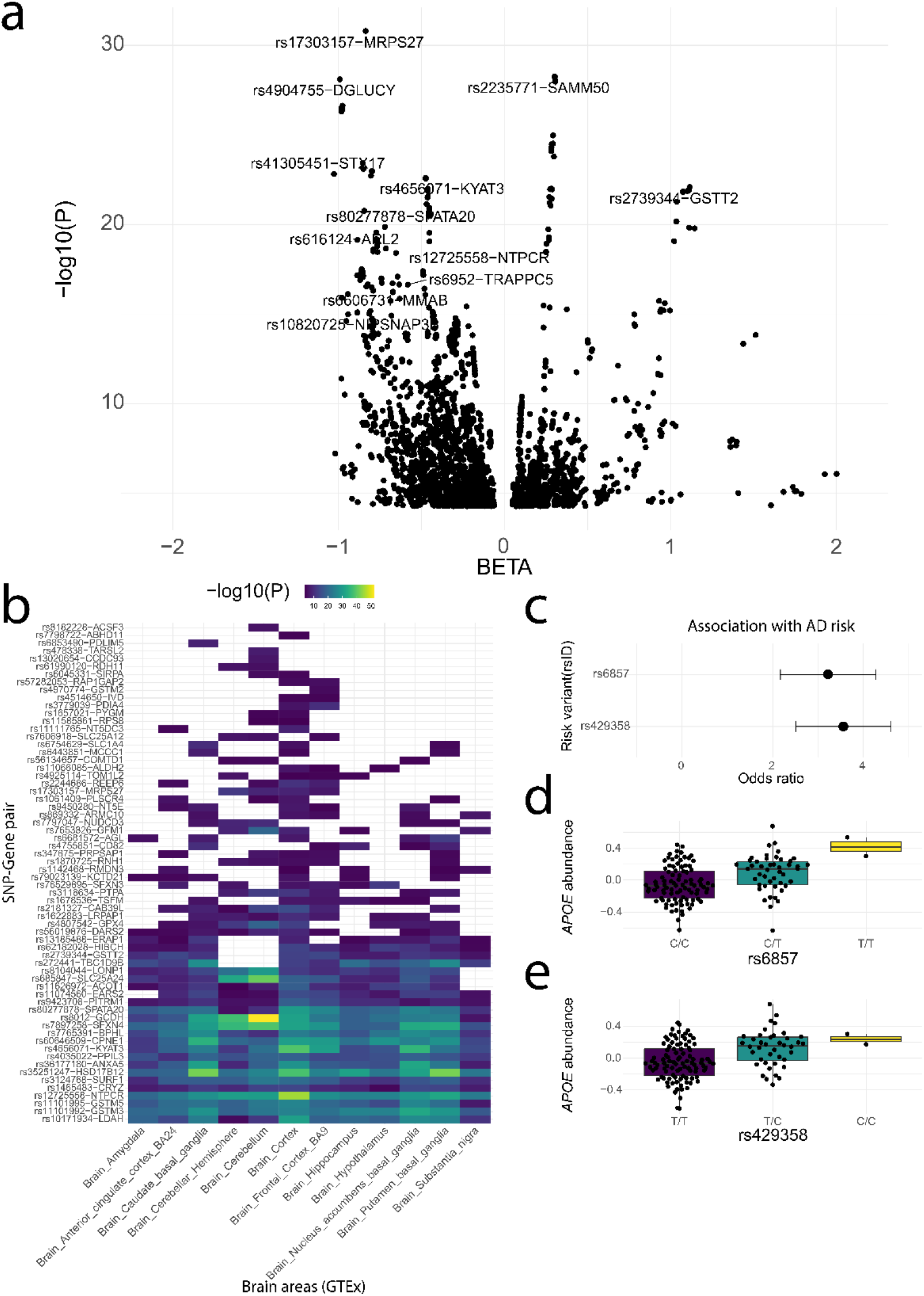
Overview of found significant pQTLs. a) Volcano plot of significant pQTLs (P_FDR_ ≤ 0.05). X-axis represents the effect size of the pQTL association, and the y-axis represents -log_10_ P-value of the pQTL association. Labels are shown for pQTLs with P-value ≤ 1 × 10^-15^. b) Heatmap of all significant pQTL variants that are also an eQTL variant. Colored squares indicate significant eQTLs based on the GTEx database, with color representing the -log_10_ P-value. The x-axis shows the tested brain tissues, and the y-axis the protein/gene names. c) Association of variants with AD risk. The x-axis represents the odd ratio (with error bars), and the y-axis shows the two significant variants. d,e) Boxplots of APOE protein residuals grouped by genotypes of d) rs6857, and e) rs429358. The x-axis represents the genotypes of rs6857/rs429358, and the y-axis represents the residuals of APOE protein residuals.

Next, we identified a set of 222 independent pQTL variants by prioritizing the most significant protein-variant pair within a linkage disequilibrium (LD) block (500 Kbp, R2>0.001). 64 of these 222 pQTL variants (29%) were also eQTL variants in at least one of the twelve brain areas available in the GTEx databasel^7^(Supplementary Table 2), and sixteen pQTLs variants being eQTL variants in all twelve brain areas (Fig. 2b). For the matching pQTLs and eQTLs, the direction of change in protein levels corresponded with the direction of change in gene transcript levels (Supplementary Fig. 1). Additionally, the effect sizes of matching pQTLs and eQTLs were significantly correlated across all brain regions (P ≤ 6.39 × 10^-3^, r ≥ 0.46).

This indicates that while the genetic regulation of protein abundance often operates independently of genetic regulation of mRNA levels, cases where shared modulation occurs suggest that mRNA levels serve as a true intermediate in driving differences in protein expression. In such instances, the genetic variant appears to exert its effect on protein abundance indirectly, by first altering mRNA levels. This highlights the critical role of mRNA as a mediator in specific instances of genetic influence, reinforcing the importance of examining both mRNA and protein data to fully understand the molecular mechanisms underlying these associations.

### rs429358 and rs6857 associate with increased APOE abundance and increased Alzheimer’s risk

Next, to elucidate the potential connection between AD-genetic risk factors and dysregulation of protein abundance, we investigated whether the identified pQTLs are associated with AD risk. Using an independent dataset of individuals with genetic data, which did not include any of the individuals used for the identification of pQTL associations^12–16^ (N = 6,479, N cases = 2,361 , N controls = 4,118 see Methods), we checked whether the identified 8,081 pQTL variants were associated with AD risk. We found only significant associations with AD for rs6857 and rs429358 (Fig. 2c, Supplementary Fig. 3). Rs6857 had an odds-ratio (OR) of 3.22 (P_FDR_ = 6.0 × 10^-145^) and rs429358 had an OR of 3.56 (P_FDR_ = 1.5 × 10^-167^) for AD risk. Both variants were pQTLs associated with *APOE* abundance. Rs6857 had a β of 0.18 (SE = 0.04, P_FDR_ = 2.57 × 10^-5^, Fig. 2d) and rs429358 had a β of 0.18 (SE = 0.04, P_FDR_ = 2.80 × 10^-5^, Fig. 2e) associated with APOE abundances.

**Figure 3.**
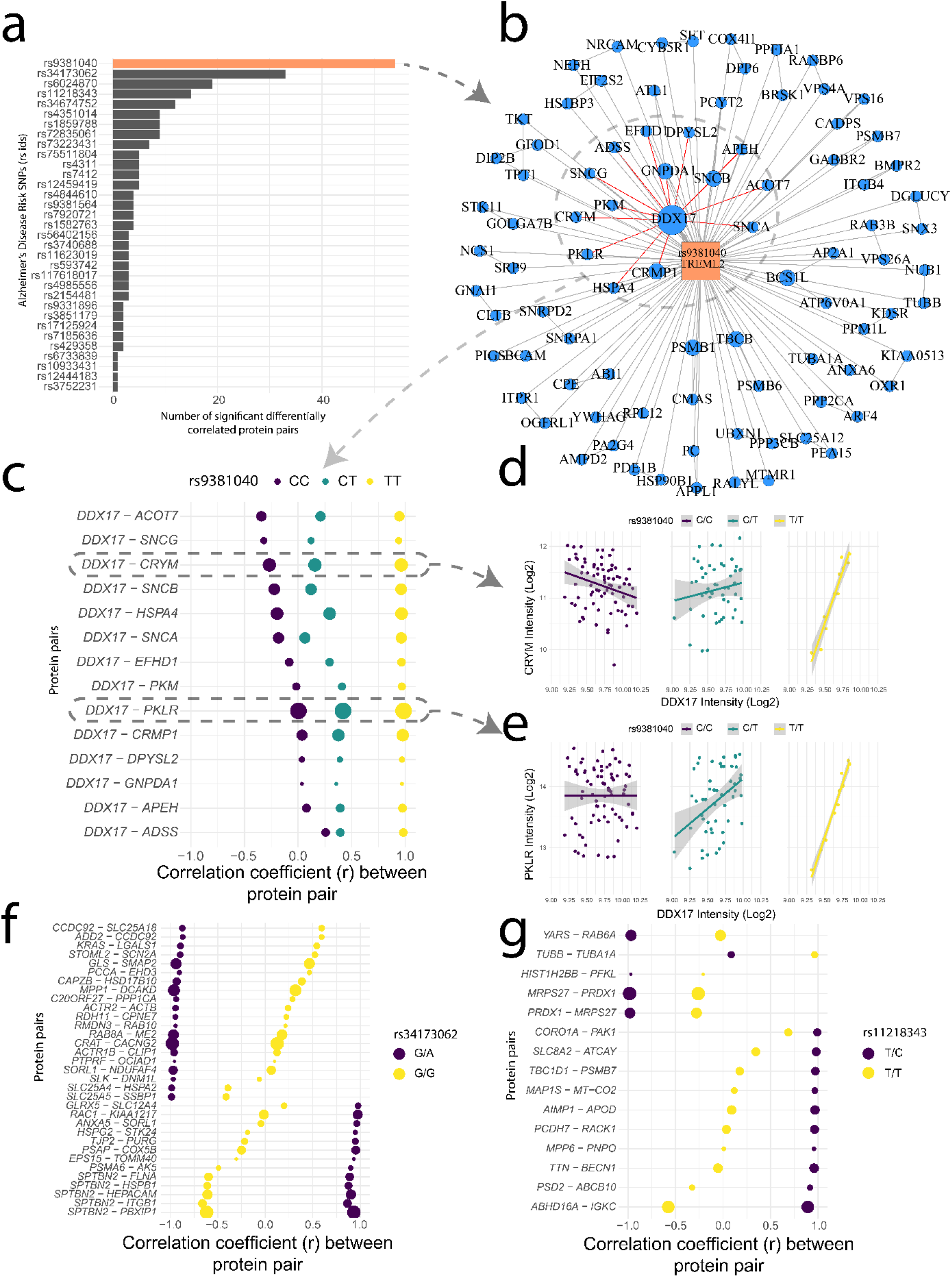
Overview of dcQTL results with respect to AD variants. a) Network graph illustrating differentially correlated proteins associated with rs9381040. Blue nodes are proteins, an edge indicates that two proteins are differentially correlated with respect to rs9381040. Node size reflects the degree of connectivity. b,c) DDX17’s differential correlation with other proteins relative to the rs9381040 genotype. The x-axis shows Pearson’s correlation coefficients between protein pairs across individuals within the respective genotypes. The y-axis lists each protein pair, and dot color indicates different genotypes. Dot size corresponds to -log_10_ p-value d, e) Scatter plots depicting two proteins (d, CRYM; e, PKLR) differentially correlated with DDX17 (x-axis) concerning rs9381040 genotypes. Each dot represents an individual, colored by genotype (purple for C/C, green for C/T, yellow for and T/T). f, g) All proteins differentially correlated with rs34173062(f) and rs11218343 (g). The x-axis represents Pearson’s correlation coefficients between the protein pairs across individuals withing the respective genotypes. The y-axis lists each protein pair, with dot colors indicating different genotypes. Dot size corresponds to -log_10_ p-value.

A Bayesian test of colocalization (see Methods *Colocalization analysis*) revealed a posterior probability (PP) of 77% that *APOE* abundance and AD risk share the same causal variant, with rs429358 being the most likely variant (PP = 1). Notably, the rs429358-T variant encodes for arginine, resulting in the APOE4 allele^17^, which is known to increase AD risk. Our findings show that the rs429358-T variant is linked to higher APOE levels in the GTM, a result not observed in prior pQTL studies of the prefrontal cortex. This may help explain the greater vulnerability of the GTM to AD, as elevated APOE4 levels could potentially lead to increased amyloid-beta accumulation specifically in this brain area.

### Genetic risk variants for Alzheimer’s Disease associate with changing associations between proteins

In addition to examining whether genetic risk variants affect the abundance of individual proteins, we explored whether AD risk variants influence the correlation between proteins, potentially indicating a genetic basis for coordinated changes in their functional associations. To investigate this, we tested for differential correlation between pairs of proteins with respect to the genotypes of 33 known AD risk variants^2^. These variants were selected such that each genotype was represented by at least 10 individuals to mitigate population size discrepancies and reduce the likelihood of false positives (Supplementary Table 3). Differential correlation QTL (dcQTL) analysis (see Methods) was performed on the 140 individuals for whom both genetic and proteomics data were available. For each of the 33 variants, 6,320,790 test were performed, testing each possible protein pair. We identified 238 pairs of proteins that were significantly differentially correlated with respect to one of the 33 AD risk variants at significance threshold P_FDR_< 0.05 (P<5.64x 0^-8^, Supplementary **Table 4**).

To assess whether the identified protein pairs could have a functional association, we checked if the protein pairs were expressed in the same cell type (FPKM > 0.1, See Methods *Human brain cell type transcriptome profile*). All protein pairs shared cell types in which they were expressed, except for two pairs (*DDX17*–*PKLR* and *HNRNPL*-*ICAM5*), which could not be validated as *PKLR* and *ICAM5* did not exceed the expression threshold in any cell type. These findings reveal that although most AD risk variants do not significantly impact the abundance of individual proteins in the GTM, they are associated with modifications in protein-protein correlation patterns.

### Potential role for DDX17 in mediating the protective effect of rs9381040-T through tight regulation of synuclein abundance

Next, we aimed to characterize the risk variants and their respective proteins whose associations with other proteins were altered in relation to the respective AD risk variants. We found that three SNPs (rs9381040 in TREML2, rs34173062 in SHARPIN and rs11218343 near SORL1) were associated with the most altered protein-protein correlations. Specifically, most differentially correlated proteins (23%) were found between the two homozygous genotypes of variant rs9381040 (closest gene = TREML2, 54 differentially correlated pairs, including 90 different proteins, Fig. 3a). These 90 proteins were enriched for neuron cell type markers (N = 17, OR = 3.55, 95%CI = 1.47-9.17, P = 2.71 × 10^-3^), suggesting that rs9381040 may predominantly influence neuronal protein networks and potentially impact neuronal function in AD.

Among the 54 differentially correlated protein pairs, DDX17 had the most altered associations, being differentially correlated with 15 proteins (Fig 3b-d). Functional enrichment analysis of the proteins differentially correlated with DDX17 revealed enrichment for ‘PFAM Protein Domains: Synuclein’ (N = 3, 1.38 × 10^-7^). All members of synuclein family (SNCA, SNCB and SNCG, Δr ≥ 1.15, P_FDR_ ≤ 2.98 × 10^-2^) were differentially correlated with DDX17 with respect to the genotypes of rs9381040. In individuals with the T/T genotype (protective), DDX17 was highly correlated with SNCA, SNCB and SNCG (r ≥ 0.94). These results suggest that DDX17 may play a protective role in individuals with the rs9381040-T/T genotype by regulating synuclein levels, potentially preventing its aggregation and the subsequent neurotoxicity seen in AD^18,19^.

33 pairs of proteins (14%, including 60 different proteins, Fig. 3c, Fig. 3d) showed differential correlation between homozygous rs34173062-G and heterozygous rs34173062-G/A. Rs34173062 is a missense variant in SHARPIN. Among these 33 differentially correlated protein pairs, SPTBN2 exhibited the most altered associations with other proteins (N=5). SPTBN2, a brain spectrin, has been implicated in several neurodegenerative diseases^20,21^, including AD^22,23^.

These findings suggest a potential association between SHARPIN and SPTBN2 concerning rs34173062. However, the functional association of this association to AD risk requires further investigation. The set of 60 proteins was not enriched for specific cell type markers. 32 of these proteins were involved in GO biological process of ‘transport’ (P_FDR_ = 8.54 × 10^-5^), 50 proteins were associated with the GO cellular component ‘cytoplasmic part’ (P_FDR_ = 5.89 × 10^-6^), and 16 proteins were linked to the mitochondrial part (P_FDR_ = 7.03 × 10^-6^).

We observed that 15 protein pairs were dcQTL with variant rs11218343 (closest gene = *SORL1*, involving 29 proteins, Fig. 3e, Fig. 3f). These proteins showed distinct correlations between homozygous (T/T) and heterozygous (T/C) individuals. However, the group of 29 proteins did not show enrichment for specific cell type markers. 18 of these proteins were involved in the *GO* biological process of ‘localization’ (P_FDR_ = 4.90 × 10^-3^), 8 proteins were associated with the *GO* cellular component ‘dendrite’ (P_FDR_ = 1.00 × 10^-4^), and 9 proteins were linked to ‘neuron projection’ (P_FDR_ = 6.80 × 10^-4^).

## Discussion

We conducted a comprehensive pQTL analysis in the GTM, identifying associations between the expression of 227 proteins and 8,081 genetic variants. Our findings align with previous studies in proteomics and transcriptomics to a certain degree, but also revealed novel variant-protein associations that may indicate specific vulnerabilities of brain regions to AD. While not many AD variants associate with differential abundance of a single protein, our results suggest that many do associate with altered associations between proteins. Specifically, our comparison of AD genetics with brain proteomics suggests that individuals carrying AD-associated variants in/near *TREML2, SHARPIN* and *SORL1* exhibit distinct protein-protein correlation structures compared to non-carriers. This implies that individual AD associated risk alleles may exert significant control over protein-protein correlation patterns, highlighting potential mechanisms underlying AD pathogenesis.

The identified dcQTLs often involved central *hub* proteins. For example, variant rs9381040 (near *TREML2* gene) was associated with differential correlations among 54 pairs of proteins. In this network, *DDX17* (DEAD-Box Helicase 17) played a pivotal role by exhibiting differential correlation with 15 proteins. *DDX17* functions as a transcriptional coregulator for various target genes^24^ and plays a significant role in the androgen signaling pathway^24^, which is implicated in protective mechanisms against neurodegenerative diseases by potentially reducing β-amyloid accumulation^25^.

Moreover, *DDX17* is involved in amyloidogenesis, a crucial process linked to AD pathogenesis, and a possible mediator contributing to AD^26^. Although *TREML2* (Triggering Receptor Expressed On Myeloid Cells Like 2) was not measured in our proteomics dataset, its expression is known to increase in neutrophils and macrophages during immune responses to inflammatory factors^27^. We speculate that the protective effects observed with rs9381040 may involve altered functions of DDX17 and potentially be initiated through immune-related responses mediated by TREML2. However, further research is needed to elucidate the functional relationship between *TREML2* and *DDX17* regarding their protective action against AD.

*SPTBN2* (Spectrin Beta, Non-Erythrocytic 2) was identified as *hub protein* linked to the effects of rs34173062, a missense variant within *SHARPIN*. Brain spectrins have garnered attention in various neurodegenerative diseases^20,21^, including AD^22,23^. Our findings suggest a connection between *SHARPIN* and *SPTBN2* concerning genetic risk for AD. The functional implications of this association in relation to AD risk require further investigation.

QTL studies traditionally aim to uncover how genetic variations regulate transcription or protein levels. However, our findings underscore the importance of moving beyond straightforward associations between genetic variants and biomolecules. A variant may not directly influence the expression levels of a transcript or protein; instead, it could trigger changes in biological states where the interactions between proteins or transcripts are redefined. While studies using single-cell RNA sequencing data increasingly adopt this perspective^28,29^, our research highlights the continued relevance of bulk data, particularly in proteomics, for unraveling these intricate genetic interactions.

This study benefits from the inclusion of single cell data to complement bulk data-derived results. By considering cell type specificities, we verified that nearly all differentially correlated proteins were expressed in the same cells, thereby affirming the hypothesis of functional association through co-expression in our findings. Another strength lies in the inclusion of individuals with extreme phenotypes, particularly cognitively healthy centenarians, who were found to be depleted with genetic variants associated with increased AD risk. This inclusion enhances the power and effect size for AD specific variants^30^.

However, it’s important to note that applying our differential correlation approach on a proteome- and genome-wide scale is impractical due to the vast number of pair-wise tests between proteins and variants involved. Therefore, a hypothesis driven approach, as employed in this study, becomes essential.

In summary, our study bridged genetic variants with proteomics in the GTM. Moreover, through differential correlation analysis based on genotypes of AD risk variants, we demonstrated that this approach holds promise as a valuable addition to GWAS- and QTL-studies. It effectively identifies proteins (e.g. DDX1 and *SPTBN2*) potentially involved in downstream effects of disease-associated risk variants. Additionally, our findings present promising results and suggest new research opportunities for exploring genetic implications of AD risk variants.

## Online methods

### Population of the study

Individuals classified as AD patients were derived from two sources: 1) clinically diagnosed with probable AD patients from the Amsterdam Dementia Cohort^12^ (N=2,668), and 2) pathologically confirmed AD patients from the Netherlands Brain Bank^14^ (N=436). The non-demented controls included individuals from various cohorts: 1) 1,779 individuals aged 55-58 years from the Longitudinal Aging Study Amsterdam^31^ (LASA), 2) 1,206 individuals with subjective cognitive decline assessed at the memory clinic of the Alzheimer center Amsterdam confirmed as cognitively normal after thorough examination, 3) 40 healthy individuals from the Netherlands Brain Bank, 4) 201 individuals from the twin study^15^, and 5) 444 individuals from the 100-plus Study cohort^16^. The 100-plus Study cohort comprises of Dutch-speaking individuals aged 100 years and older, who self-reported to be cognitively healthy confirmed by their family members and partners. For this study, a total of N=358 cognitive health centenarians and N=86 partners of centenarian’s children were considered. Genetic data was available for all individuals. The Medical Ethics Committee of the Amsterdam UMC (METC) approved all studies. All participants and/or their legal representatives provided written informed consent for participation in clinical and genetic studies.

### Genetic data processing

Genetic variants were identified using standard genotyping and imputation methods, followed by established quality control procedures. Genotyping was performed on individuals using Illumina Global Screening Array (GSAsharedCUSTOM_20018389_A2). High-quality genotyping was retained (individual call rate > 98%, variant call rate > 98%), with exclusions for sex mismatches and significant departures from Hardy–Weinberg equilibrium (P<1×10^-6^). Genotypes were prepared for imputation using provided scripts (HRC-1000G-check-bim.pl)^32^ to compare variant ID, strand and allele frequencies to the Haplotype Reference Panel (HRC v1.1, April 2016)^33^. All autosomal variants were submitted to the Sanger imputation server (https://imputation.sanger.ac.uk), which uses MACH for phasing and PBWT for imputation against the HRC v1.1, April 2016 reference panel.

In total, 3,670 population subjects and 3,106 AD cases passed quality control. Before analysis, individuals of non-European ancestry were excluded based on 1000Genomes clustering, those with a family relation (identity-by-descent > 0.2) were also excluded. This resulted in the exclusion of 205 population controls and 152 AD cases with non-European ancestry, and 217 population controls and 100 AD cases with family relations. Consequently, 4,191 control subjects and 2,416 AD cases remained for the analyses, yielding a total sample size of 6,607. Of these, 140 individuals also had proteomics data.

### Summary statistics pQTL study

The pQTL summary statistics of Robins et al.^8^ were obtained from http://brainqtl.org. The pQTLs were identified in 144 healthy individuals originally part of the ROSMAP study, a population composition of 63.1% females and a median age of 86.5 (range: 67.4 to 102.7). Protein expression data were sourced from the dorsolateral prefrontal cortex. In contrast to our pQTL analysis, variants within 50 Kbp up- and downstream of the TSS of the respective proteins were tested. The dataset includes pQTL summary statistics for 7,901 proteins and 2,599,383 variants, totaling, 4,199,577 pQTLs. Proteins were identified using their Uniprot accession IDs and variants by their GRCh37/hg19 genomic coordinate. P-values were corrected for multiple tests using both Bonferroni and FDR methods. Using Bonferroni correction, 2,955 significant pQTLs (P_BONF_ ≤ 0.05) were identified, while FDR correction identified 28,211 significant pQTLs (P_FDR_≤ 0.05).

### eQTLs from GTEx

The eQTL data was accessed through the GTEx Application programming interface (API)^7^. Twelve brain regions were analyzed, each brain with varying numbers of individuals who had genotype and RNA-seq data available (Table S1). The total population comprised 395 individuals, 72% of whom were male. While GTEx does not report the exact ages of these individuals, the specified age ranges are as follows: 20-29 years (N = 8), 30-39 years (N = 10), 40-49 years (N = 36), 50-59 years (N = 119), 60-69 years (N = 200), and 70-79 years (N = 22).

The eQTL statistics obtained from GTEx include a normalized effect size (NES), which is the slope of the linear regression, indicating the effect of the alternative allele (ALT) relative to the reference allele (REF) according to human genome reference GRCh38/hg38. The data also contains nominal p-values for the eQTL association and a p-value threshold, determined by P_FDR_ ≤ 0.05, but is translated to a nominal p-value. Variants are defined by their reference SNP identification number (rs IDs), while transcripts are defined by their gene symbol and an Ensembl transcript ID.

### Gyrus Temporalis Medialis Proteomics data

Proteomics data from the GTM was measured for a total of 237 individuals from Netherlands Brain Bank^34^, from whom measurements were taken for 3,555 proteins. Among them, 102 individuals were diagnosed with Alzheimer’s Disease (AD), 62 were cognitively health centenarians (CHC), and 73 were non-demented (ND) controls. Proteomics data was generated using the Sequential Window Acquisition of All Theoretical Mass Spectra (SWATH-MS) method employing a Data-Independent Acquisition (DIA) approach. Spectrum annotation and relative protein quantification were performed using MaxQuant software^35^, with the Uniprot human reference proteome^36^ used as reference.

### Gyrus Temporalis Medialis Proteomics Quality Control and Pre-processing

Quality control was performed separately on both a sample basis and protein basis. Initially, samples with more than 34% of low-quality peptides (Q ≥ 0.01) were excluded from the analyses (N = 35). After removing low-quality samples, a reference peptide intensity distribution was calculated by averaging the peptide intensity distributions of the remaining samples. The distance between each individual peptide intensity distribution and the reference distribution was then calculated using the Kolmogorov–Smirnov test. Samples with a distribution distance (*D*) greater than 0.04 from the reference distribution were excluded from the analyses (N = 1). For replicate samples, lower quality samples were determined using a paired t-test on the quality measures, resulting in the exclusion of eleven replicates.

The proteomics data was generated in bottom-up fashion, where peptides were measured and used to estimate the expression of their respective protein. If the peptides comprising a single protein were of low-quality in more than 10% of the samples, the respective protein was excluded. Protein amount were represented by the sum of intensities of their respective peptides. Finally, protein intensities were log_2_ transformed to ensure the normality of the protein intensity distributions. The final proteomics dataset consists of 3,555 proteins and 190 individuals.

Next, batch effects, which have no biological meaning, were removed during the pre-processing step. Initially, the association between variations in the proteomics data and the variables age, sex, Braak stage I-VI, post-mortem delay (PMD), *APOE* genotype (log_2_ Polygenic Risk Score), and batch were tested using the R-package variancePartition^37,38^. VariancePartition employs a mixed linear model to determine the percentage of variation attributable to each variable. Among the tested variables, substantial proportions of the variation were explained by age, Braak stage and batch. To remove the variation associated with batch from the protein intensity data, the combat function from the R-package sva^39^ was used.

### Human brain cell type transcriptome profile (HBCT)

The cell type specific transcript expression data of Zhang et al.^40^ were obtained from https://www.brainrnaseq.org/. A total of 21.661 transcripts were measured across five cell types: astrocytes, neurons, oligodendrocytes, microglia/macrophages, and endothelial cells, sampled from individuals ranging from 8 years to 63 years old. Specifically, transcripts measurements in astrocytes were derived from 26 samples, with fourteen from male individuals and twelve from females, including four samples from the tumor core or their vicinity. Neuron-specific expression came from one male individual, while oligodendrocyte-specific expression was derived from 5 individuals (4 makes and 1 female). Microglia/macrophages expression was available from three individuals (2 males and 1 female), and the endothelial cells from two females. The transcript expression levels were normalized using Fragments Per Kilobase Million (FPKM), providing a standardized representation of transcript abundance.

### Cell type markers, composition, and enrichment

Cell type markers were estimated using the HBCT transcriptome dataset. First, cells that originated from a tumor or its surroundings were excluded (N = 4). The average FPKM of each gene, in each of the five cell types (astrocytes, neurons, oligodendrocytes, microglia/macrophages, and endothelial cells) was calculated. For genes with multiple measurements, an average FPKM value was calculated. A gene was annotated as unique cell type marker for a particular cell type when the fold change of the average FPKM was ≥ 3 compared to all other cell types.

Subsequently, the cell type composition for individuals with available protein data (N = 190, See Table 1: *Protein data*) was estimated. This was achieved by averaging the protein intensities of the unique cell type makers present in the GTM protein dataset for each cell type, thus providing an estimation of the cell type abundance for each individual based on their respective protein abundance. Finally, cell type enrichments were performed with the Fisher’s exact test^41^, based on the unique cell type markers of the five cell types, to calculate the enrichment of a set of proteins for unique cell type markers.

### pQTL identification

pQTL analysis was performed on the subset of 140 individuals for which both genetics and proteomics data was available after quality control. Genetic variants associated with protein expression were identified with Plink (v2.00a2LM)^42^. Linear models were employed for the association analysis, with genotype dosages as predictors for protein expression, assuming additive genetic effects. The analyses were corrected for estimated cell type composition (neurons, microglia/macrophages and oligodendrocytes, See Materials and methods *Cell type markers, composition, and enrichment*) and population substructure using the first five principal components (See methods *pQTL linear regression model*). An additional analysis was performed correcting for phenotype status (AD, ND controls and CHC, See Table 1: *intersection*).

Resulting effect-sizes (β) were calculated with the minor allele relative to the major allele in our population. Association P-values were corrected for multiple tests with False Discovery Rate (FDR), with assumed at P_FDR_ ≤ 0.05. The analyses were restricted to variants with a minor allele frequency (MAF) higher than 5% and variants located 250 Kbp down- and upstream of the TSS of the respective proteins. Four window sizes were tested (50 Kbp, 250 Kbp, 500 Kbp and 1 Mb, Supplementary Fig. 4). Using the 250 Kbp window, we reduced to total number of tests while still capturing most of pQTLs.

Genomic locations of the TSSs were acquired with biomaRt (v2.42.0)^43,44^. The retrieved genomic locations of the TSSs were for genomic build GRCh38/hg38. The liftOver R-package (v1.10.0)^45,46^ was used to lift over the genomic coordinates to build GRCh37/hg19, as the genotype files were based on this genomic build.

### pQTL linear regression model

For identifying significant pQTLs, the generalized linear model (GLM) from Plink (v2.00a2LM)^42^ was used, which is the primary association analysis method in Plink for quantitative phenotypes. The model applied to our data was as follows:

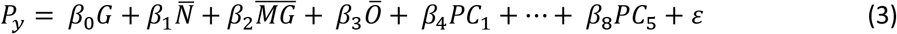

where: P_y_ is the log_2_ intensity for the individuals of respective protein (quantitative phenotype); G are the dosages for the individuals of the respective variant that is tested; 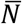 are the mean intensity of all Neuron cell type markers;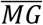 are the mean intensity of all Microglia/Macrophage cell type markers;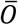 are the mean intensity of all Oligodendrocytes cell type markers; PC_i_ are the principal components for the individuals of the population substructure; *ε* is an error term that gets minimized with least squares minimization.

### Testing pQTL variants on association with AD risk

All significant pQTL variants were tested on association with AD risk using all individuals for which genetics data was available. This, which comprised 6,479 individuals (2,361 AD cases and 4,118 ND controls). To ensure an independent population, individuals used in the pQTL identification (N=128 of 140) were excluded, ensuring no overlap of individuals between the differential expression analysis population and the genetic association test population. The association of pQTL variants with AD status was tested using a logistic regression model in R (v3.6.3), with AD status as discrete outcome variable (ND = 0, AD = 1) and the pQTL variant’s dosages as predictor variable. The model was adjusted for population substructure using the first five principal components. P-values were adjusted for multiple tests using FDR, with significant association assumed at P_FDR_ ≤ 0.05.

### Colocalization analysis

For the colocalization analysis, we initially identified proteins associated with pQTL variant that also showed associated with AD status (See methods *Testing pQTL variants on association with AD risk*). Subsequently, we extracted all variants within 250 Kbp up- and downstream of the TSS of these proteins. These variants were then evaluated for association with AD status using the same cohort of individuals described previously (N = 6,479, See methods *Testing pQTL variants on association with AD risk*).

A logistic regression model was performed with AD status (ND = 0, AD = 1) as the discrete outcome variable and genotypes of the aforementioned variants as predictor variable. The model was adjusted for population substructure using the first five principal components.

To estimate the probability that the specified genomic region contains a pQTL variant influencing both protein abundance and AD risk, an approximate Bayes Factor colocalization analysis was performed using the coloc R-package (v3.2.1)^47^. This analysis utilized summary statistics including p-values, sample size and MAF. The colocalization analysis tests five hypotheses. *H*_*0*_: There is no association of the genomic region with protein abundance and AD risk. *H*_*1*_: There is only an association with protein abundance. *H*_*2*_, there is only an association with AD risk. *H*_*3*:_ The genomic region is associated with both protein abundance and AD risk, but through two different variants. *H*_*4*_: The genomic region is associated with both protein abundance and AD risk, through a single variant. For each hypothesis, a posterior probability is computed to assess the likelihood of colocalization between protein abundance and AD risk in the specified genomic region.

### pQTL and eQTL comparison

We examined whether the significant pQTLs were also an eQTL variant using the eQTL data from twelve brain tissues (Table S1) from GTEx (v8)^7^. An independent set of clumped pQTLs (See methods *Clumping)* was used to minimize the number of requests that needed to be sent to the API of GTEx. We investigated associations of pQTLs from the GTM with eQTLs both across the twelve brain tissues and within specific brain regions. Each gene-variant pair was queried across twelve brain tissues using the get_eQTL_bulk function from the R-package CONQUER (v1.0)^48^, which requires tissue ID, gene symbol and RS ID to be supplied. Significance of the tested eQTLs was determined using the P-value thresholds provided by GTEx. To compare the directional effects of pQTLs with their synonymous eQTLs, we calculated Pearson’s correlation coefficient between effect sizes using the cor.test function.

### Clumping

We created an independent set of pQTLs using LD-based clumping. Variants that are located close to each other often exhibit linkage disequilibrium, meaning that they are correlated and show similar associations with the same protein. The clumping procedure allows to retain only the most strongly associated variant within a specified window. We performed clumped individually for each protein using Plink (v1.90b4.6)^42^, with criteria set at R^2^ ≥ 0.001 and MAF ≥ 0.05. Linkage disequilibrium between variants was calculated using European individuals from the 1,000 Genomes Project reference panel^49^.

### Differential correlation

We examined whether protein correlation changed when comparing two distinct groups based on AD variant genotypes. Initially, Pearson’s correlation between a pair of proteins was calculated separately for each group of interest. Let’s denote these coefficients as *r*_*x*_ for group *x* and *r*_*y*_ for group *y*. Subsequently, these correlation coefficients were transformed into z-scores using Fisher’s z-transformation^50^ (Eq. 4).

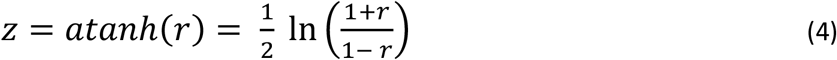

The difference between z-scores *z*_*x*_ and *z*_*y*_ was then calculated using equation 5:

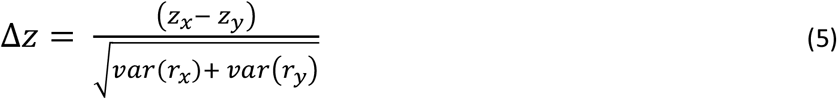

where *var*(*r*) is calculated by 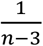 , with *n* being the sample size of the respective groups. Since Δ*z* follows a normal distribution, a two-sided P-value for the differential correlation between each pair of proteins can be determined.

### Differential correlation with respect to AD variants genotype

In this analysis, we used individuals with both genetics and proteomics data: 67 AD individuals, 27 ND controls, and 46 CHC. We conducted a differential correlation analysis of proteins based on the genotypes of established AD variants. Initially, we considered 41 variants^2^ known to influence AD risk. We selected variants where each genotype was represented by at least 10 individuals to mitigate population size discrepancies and reduce the likelihood of false positives, resulting in 33 remaining variants (Supplementary Table 3).

For variants where all three genotypes were present, we calculated the differential correlation between the two homozygous genotypes, reporting the correlation involving the heterozygous genotype separately. When only two genotypes were present, we assessed the differential correlation between the homozygous genotype and the heterozygous genotype.

The differential correlation method for these variants was implemented in R (v3.6.3)^51^. P-values were FDR corrected based on the total number of tests conducted. Significance was established at P_FDR_ ≤ 0.05.

## Data Availability

All data produced in the present study are available upon reasonable request to the authors.

