## Supplementary file for "Genetic Variants that Modulate Alzheimer’s Disease Risk Deregulate Protein-Protein Correlations in the Gyrus Temporalis Medius"

### Supplements

***pQTL linear regression model***

For identifying significant pQTLs, the generalized linear model (GLM) from Plink (v2.00a2LM)[60] is used. Which is the primary association analysis method in Plink for quantitative phenotypes. The model applied on our data was as follows:

$$P_y = \beta_0 G + \beta_1 \overline{N} + \beta_2 \overline{MG} + \beta_3 \overline{O} + \beta_4 PC_1 + \cdots + \beta_8 PC_5 + e$$

Where:

$P_y$ is the $\log_2$ intensity for the individuals of respective protein (quantitative phenotype).

G are the dosages for the individuals of the respective variant that is tested.

$\overline{N}$ are the mean intensity of all Neuron cell type markers

$\overline{MG}$ are the mean intensity of all Microglia/Macrophage cell type markers

$\overline{O}$ are the mean intensity of all Oligodendrocytes cell type markers

$PC_i$ are the principal components for the individuals of the population substructure.

e is an error term that gets minimized with least squares minimization.

#### Protein residuals

For each individual, for each protein measured in the GTM protein dataset we calculated the residual after correcting for the abundance of three cell types (neurons, microglia/macrophages, and oligodendrocytes). The pQTL analysis was also corrected for the abundance of these cell types. The residuals were calculated in order to truthfully visualize the pQTL associations.

First for each protein we fitted a linear model:

$$Protein_{int} = \beta_0 + \beta_1 \overline{N} + \beta_2 \overline{MG} + \beta_3 \overline{O} + \varepsilon$$

Where:

- $\overline{N}$ = the mean intensity of all neuron cell type markers
- $\overline{MG}$ = the mean intensity of all microglia/macrophage cell type markers
- $\overline{O}$ = the mean intensity of all oligodendrocytes cell type markers

Next, with the fitted model, we predicted the protein expression:

$$Protein_{pred} = \beta_0 + \beta_1\overline{N} + \beta_2\overline{MG} + \beta_3\overline{O} + \varepsilon$$

And finally, we subtracted $Protein_{pred}$ from $Protein_{int}$ to get the protein residuals.

**S Table 2: Genotyped and RNAseq sample sizes from GTEx for all twelve investigated brain regions**

| Tissue | # RNASeq and Genotyped samples | # RNASeq Samples |
|---|---|---|
| *Cerebellum* | 209 | 241 |
| *Cortex* | 205 | 255 |
| *Nucleus accumbens (basal ganglia)* | 202 | 246 |
| *Caudate (basal ganglia)* | 194 | 246 |
| *Cerebellar Hemisphere* | 175 | 215 |
| *Frontal Cortex (BA9)* | 175 | 209 |
| *Hypothalamus* | 170 | 202 |
| *Putamen (basal ganglia)* | 170 | 205 |
| *Hippocampus* | 165 | 197 |
| *Anterior cingulate cortex (BA24)* | 147 | 176 |
| *Amygdala* | 129 | 152 |
| *Substantia nigra* | 114 | 139 |

**Table S3: AD risk variants subject in differential correlation analysis**

| RS ID | Chromosome | Genomic location | Closest Gene | Genotypes | | |
|---|---|---|---|---|---|---|
| **rs6733839** | 2 | 127892810 | *BIN1* | C/C 43 | C/T 71 | T/T 26 |
| **rs9381040** | 6 | 41154650 | *TREML2* | C/C 78 | C/T 51 | T/T 11 |
| **rs1859788** | 7 | 99971834 | *PILRA* | A/A 13 | A/G 58 | G/G 69 |
| **rs73223431** | 8 | 27219987 | *PTK2B* | C/C 43 | C/T 81 | T/T 16 |
| **rs9331896** | 8 | 27467686 | *CLU* | C/C 18 | C/T 64 | 58 T/T |
| **rs34674752*** | 8 | 145154222 | *SHARPIN* | A/A 0 | G/A 11 | G/G 129 |
| **rs7920721** | 10 | 11720308 | *ECHDC3* | A/A 60 | A/G 56 | G/G 24 |
| **rs3740688** | 11 | 47380340 | *SPI1* | G/G 22 | G/T 68 | T/T 50 |

| | | | | | | |
|---|---|---|---|---|---|---|
| **rs1582763** | 11 | 60021948 | *MS4A4A* | A/A<br>18 | G/A<br>77 | G/G<br>45 |
| **rs3851179** | 11 | 85868640 | *PICALM* | C/C<br>53 | T/C<br>66 | T/T<br>21 |
| **rs11218343*** | 11 | 121435587 | *SORL1* | C/C<br>0 | T/C<br>12 | T/T<br>128 |
| **rs12444183** | 16 | 81773209 | *PLCG2* | A/A<br>20 | A/G<br>65 | G/G<br>55 |
| **rs4311** | 17 | 61560763 | *ACE* | C/C<br>34 | T/C<br>75 | T/T<br>31 |
| **rs12459419** | 19 | 51728477 | *CD33* | C/C<br>68 | C/T<br>58 | T/T<br>14 |
| **rs2154481** | 21 | 27473875 | *APP* | C/C<br>35 | C/T<br>64 | T/T<br>41 |

*** = variant of which two genotypes were present in population**

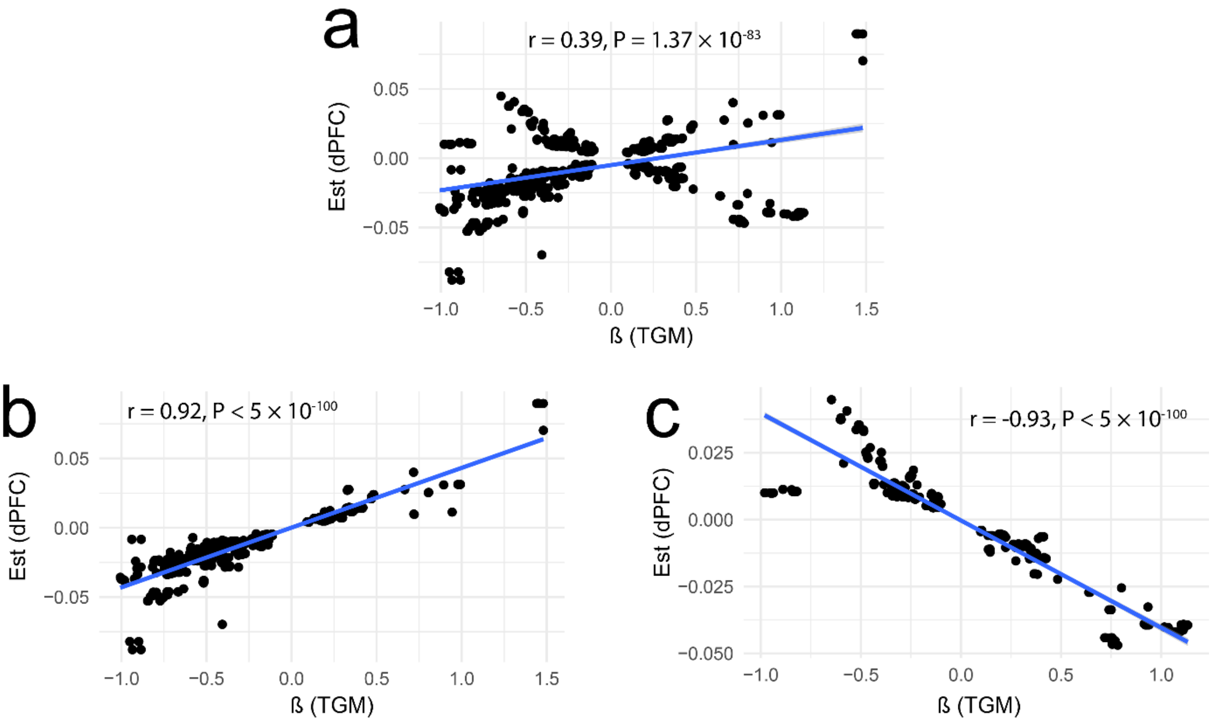


**Figure s1** Estimate comparison of pQTL studies, in all sub-figures the x-axis represents the estimates of this current study and y-axis represents the estimates from [16]. a) Estimates of all matching pQTLs. b) Estimates of all matching pQTLs where the directional effects were the same. c) Estimates of all matching pQTLs where the directional effects opposite.

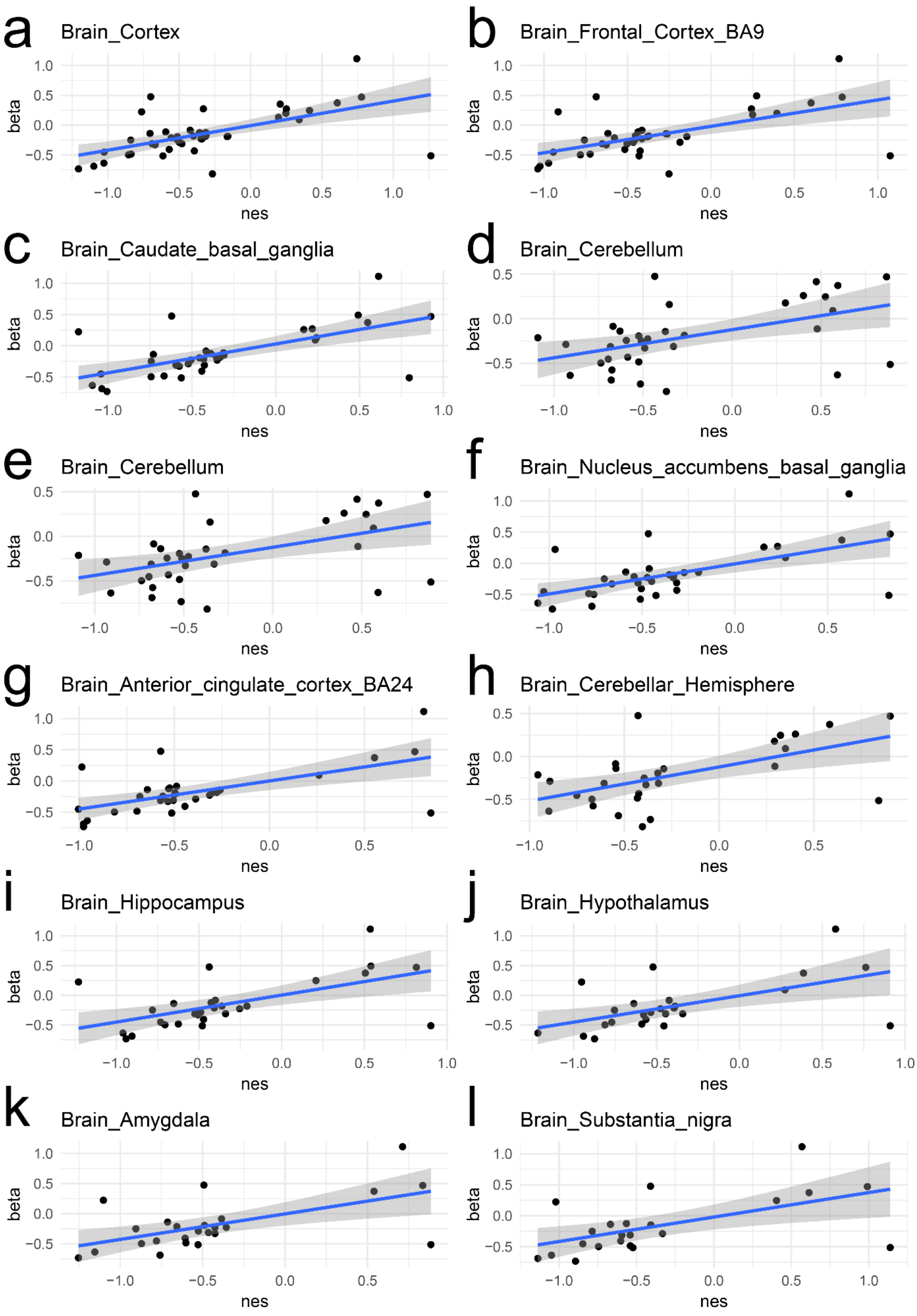


**Figure s2** Estimate comparison of pQTLs versus the eQTL NESs from GTEx for all investigated brain regions. X-axes represent the NESs from GTEx for a particular eQTL – eGene pair. The y-axes represent the betas of the pQTL – protein pair synonymous for the eQTL – eGene pair.

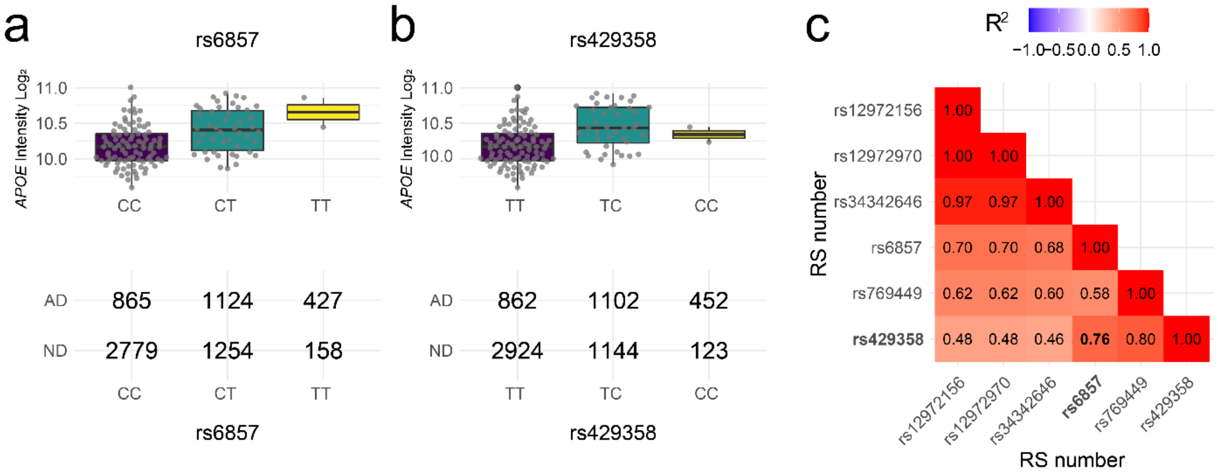


**Figure s3** Overview of APOE associated pQTL variants. a) Boxplot of rs6857 genotypes versus APOE intensity, x-axis represent the genotypes, y-axis represents the $\log_2$ normalized intensity of APOE. b) Boxplot of rs429358 genotypes versus APOE intensity, x-axis represent the genotypes, y-axis represents the $\log_2$ normalized intensity of APOE. c) LD correlation between the six pQTL variants associated with APOE.

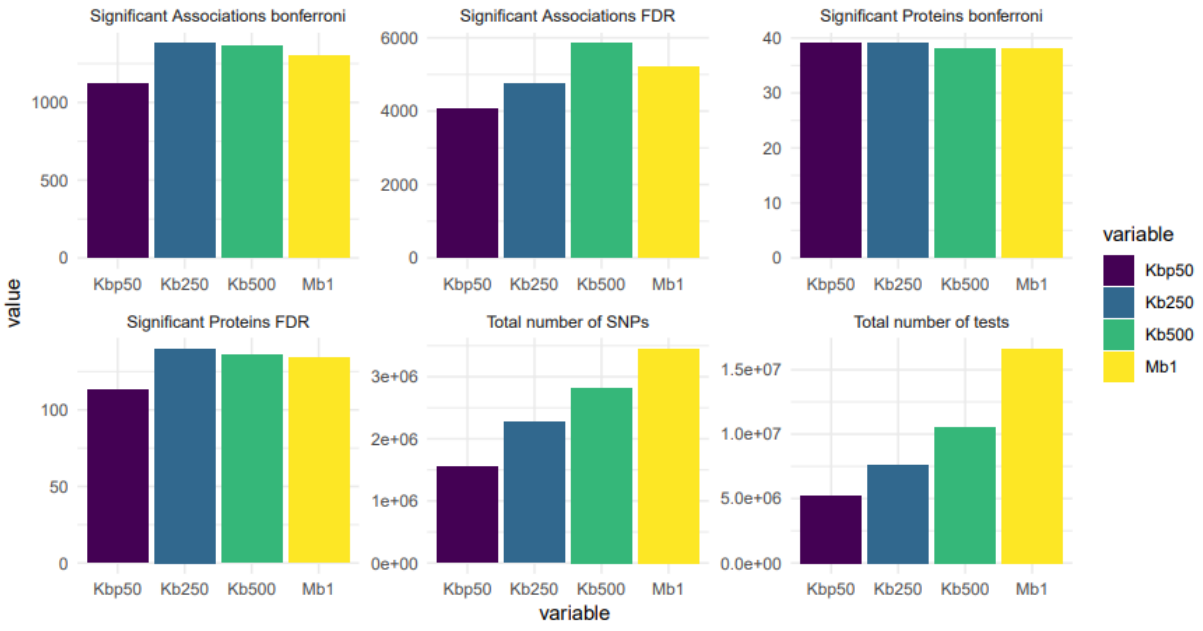


**Figure s4** Overview of pQTL variant mapping window. X-axes represent the mapping windows of 50 Kbp, 250 Kbp, 500 Kbp and 1 Mb. The y-axes represent the count of the respective statistic that is shown. The title above each plot is the respective statistic.
